# Predicting hospitalizations related to ambulatory care sensitive conditions with machine learning for population health planning: derivation and validation cohort study

**DOI:** 10.1101/2021.02.24.21252324

**Authors:** Seung Eun Yi, Vinyas Harish, Jahir M. Gutierrez, Mathieu Ravaut, Kathy Kornas, Tristan Watson, Tomi Poutanen, Marzyeh Ghassemi, Maksims Volkovs, Laura Rosella

**Affiliations:** Department of Computer Science, University of Toronto, Toronto, ON, Canada; Layer 6 AI, Toronto, ON, Canada; Dalla Lana School of Public Health, University of Toronto, Toronto, ON, Canada; Temerty Faculty of Medicine, University of Toronto, Toronto, ON, Canada; Temerty Centre for Artificial Intelligence Research and Education in Medicine, University of Toronto, Toronto, ON, Canada; School of Computer Science and Engineering, Nanyang Technological University, Singapore; ICES, Toronto, ON, Canada; Vector Institute, Toronto, ON, Canada; CIFAR AI Chair, Canada; Institute for Better Health, Trillium Health Partners, Mississauga, ON, Canada

## Abstract

**Objective:** To predict older adults’ risk of avoidable hospitalization related to ambulatory care sensitive conditions (ACSC) using machine learning applied to administrative health data of Ontario, Canada.

**Design, Setting, and Participants:** A retrospective cohort study was conducted on a large cohort of all residents covered under a single-payer system in Ontario, Canada over the period of 10 years, between 2008 and 2017. The study included 1.85 million Ontario residents between 65 and 74 years old at any time throughout the study period.

**Data sources:** Administrative health data from Ontario, Canada obtained from the ICES Data Repository.

**Main outcome measures:** Risk of hospitalizations due to ACSCs one year after the observation period.

**Results:** The study used a total of 1,854,116 patients, split into train, validation, and test sets. The ACSC incidence rates among the data points were 1.1% for all sets. The final XGBoost model achieved an AUC of 80.5% on the held-out test set, and the predictions were well-calibrated. When ranking the predictions made by the model, those at the top 5% of risk as predicted by the model captured 37.4% of those presented with an ACSC-related hospitalization. A variety of features such as the previous number of ambulatory care visits, presence of ACSC-related hospitalizations during the observation window, age, rural residence, and prescription of certain medications were contributors to the prediction. Our model was also able to capture the geospatial heterogeneity of ACSC risk in the province of Ontario, and especially the elevated risk in rural and marginalized regions.

**Conclusions:** This study aimed to predict the 1-year risk of hospitalization from a series of ambulatory-care sensitive conditions in seniors aged 65 to 74 years old with a single, large-scale machine learning model. The model shows the potential to inform population health planning and interventions to reduce the burden of ACSC-related hospitalizations.

## Introduction

Health systems globally strive to provide quality care in alignment with the Quadruple Aim [1,2]. Ambulatory Care Sensitive Conditions (ACSCs) are a useful indicator of health system performance often used to support Quadruple Aim efforts. Reducing the burden of ACSCs is an opportunity to promote higher value care by avoiding the usage of unnecessary hospital and health system resources [3]. ACSCs refer to conditions that can be managed in primary care settings, and thus hospitalization for these conditions are considered potentially avoidable with adequate primary care [4]. The list of the conditions considered to be ACSCs varies by country but commonly includes major chronic diseases [5,6,7]. For example, in Canada, ACSCs comprise angina, asthma, chronic obstructive pulmonary disease (COPD), diabetes, epilepsy, and heart failure [8]. Outcomes with respect to these ACSCs are often used to measure how a health system specifically supports individuals with chronic diseases [9]. Although only 0.4% of Canadians under the age of 75 have an ACSC-related hospitalization, these events use nearly 11% of hospital bed days [10]. In the United Kingdom, emergency admissions account for 67% of all hospital days, costing £12.5B annually, most of which are considered preventable. [11]. ACSCs have increased by 47% over the last 15 years (as of 2017) and account for one in five unplanned admissions [12]. Furthermore, older adults disproportionately bear the brunt of ACSCs. For example, adults over 65 represented 69.1% of all ACSCs in Ireland in 2016 [13] and more than half of ACSCs in Canada were from individuals over 60 years old [10]. Therefore, reducing ACSCs represent a challenge in health systems around the world and pose an increasing burden for health systems as rates of multimorbidity continue to rise [14,15].

Hospitalizations due to ACSCs are also strongly related to socioeconomic status, even in settings of universal health coverage. Studies in Canada, the UK, the US, and Ireland [16,17,18,19] have demonstrated the association between low socioeconomic status (often measured through area-level deprivation measures) and higher rates of ACSCs. As these ACSC disparities can be due to systems-level barriers, a number of community-based interventions such as pay for performance schemes or multidisciplinary programs to prevent readmissions have been proposed to potentially reduce the rates of these hospitalizations [20]. There is a need to develop tools for predicting the admissions that are most likely to be preventable to guide the deployment of such interventions or to target resources to optimize impact [20,21]. Risk prediction algorithms developed on big data sources such as electronic medical records and routinely collected administrative health data (AHD) offer the ability to segment populations over space and time based on their likelihood of having an outcome of interest [22], which can be used as an input to optimize outcomes. AHD are comprehensive, widely available, and automatically generated based on interactions with the healthcare system. Furthermore, in certain settings, these data can be linked with other data sources (e.g. environmental exposures, area-level measures of the social determinants of health, etc.) [23,24]. These characteristics make them particularly appealing for developing risk prediction algorithms that can be deployed at the level of populations for health system planning. While there is an increasing number of risk prediction models intended for clinical use in individuals, there are few examples of a single, unified model that can be deployed on routinely collected data to regularly support population health and health system management. Databases with analogous administrative health data are available in most single-payer healthcare systems such as the United Kingdom, Australia and New Zealand. However, access to extensive medical records is not limited to single-payer countries. Databases of commercial insurance claim data are also available for large portions of the population in countries with private healthcare systems such as the United States. Consequently, we believe that with sufficient adaptation, our proposed approach has wide applicability for assessing the risk of ACSCs in the population using routinely collected data.

The specific aim of this study is to develop and internally validate a single, large-scale, machine learning model to predict hospitalizations due to ACSCs in a cohort of 1.85 million older adults in Ontario, Canada. Ontario’s population is large, diverse, and covered under a single-payer system [25,26]. We extracted a wide variety of features from Ontario’s comprehensive AHD databases including socio-demographics, prescribed medications, insurance claims from physician’s visits and hospitalizations, as well as laboratory values. We evaluated model performance including discrimination, calibration, and report on the top contributing features. We finally conducted a geographical analysis to demonstrate how the model captures risk distribution in the population and thus can be used for guiding population health planning and community-based interventions.

## Methods

### Data Sources

We used administrative health data from Ontario, Canada which we obtained from ICES, an independent, non-profit research institute whose legal status under Ontario’s health information privacy law allows it to collect and analyze health care and demographic data, without consent, for health system evaluation and improvement [27]. Ontario, with a population of 14.7 million people as of April 2020 [26], is Canada’s most populous province with close to 250 reported ethnicities (2016 census [25]), making it one of the most diverse in the world. The vast majority of Ontario residents are eligible for universal health coverage, leading to highly comprehensive and representative records. Besides demographic information, the records from ICES include linked claims data that are routinely collected every time a patient interacts with the healthcare system. To link all attributes for each patient, we used a unique identification number assigned to each individual at the Registered Persons Database (RPDB). All analyses were carried out on the Health AI Data Analytics Platform (HAIDAP), which enables high performance computing in a secure environment to protect patient privacy. All datasets used from the ICES data repository are listed in Supplementary tables S1 and S2. These datasets were linked using unique encoded identifiers and analyzed at ICES.

### Cohort

Our study period spanned a 10-year window from January 1st, 2008 through December 31st, 2017. As older adults are disproportionately affected by ACSCs [10], we only included patients who were between 65 and 74 years of age at any point during the 10-year study period and who were covered under Ontario Health Insurance Plan (OHIP). Baseline characteristics of the patients included in our cohort are shown in Table 1. The methodology used to extract and prepare patient data is illustrated in Figure 1.

**Table 1.** Characteristics of the patients in the study.

| Cohort | Train |  | Validation |  | Test |  |
| --- | --- | --- | --- | --- | --- | --- |
|  | Total | Positives | Total | Positives | Total | Positives |
| Unique patients | 1,237,507 | 119,728 | 309,380 | 30,224 | 1,375,277 | 80,788 |
| Data points | 16,923,230 | 180,152 | 4,225,605 | 45,571 | 9,862,034 | 104,676 |
| **Demographics** |  |  |  |  |  |  |
| Sex (F) | 8,845,443 (52.3%) | 93,333 (51.8%) | 2,213,395 (52.4%) | 23,542 (51.7%) | 5,153,174 (52.3%) | 54,368 (51.9%) |
| Sex (M) | 8,077,787 (47.7%) | 86,819 (48.2%) | 2,012,210 (47.6%) | 22,029 (48.3%) | 4,708,860 (47.7%) | 50,308 (48.1%) |
| Age (mean) | 69.0 ± 2.86 | 69.1 ± 2.88 | 69.0 ± 2.86 | 69.1 ± 2.87 | 69.0 ± 2.81 | 69.1 ± 2.84 |
| Immigrant | 1,619,193 (9.57%) | 16,686 (9.26%) | 405,211 (9.59%) | 4,007 (8.79%) | 1,071,959 (10.9%) | 10,878 (10.4%) |
| Non-Immigrant | 15,304,037 (90.4%) | 163,466 (90.7%) | 3,820,394 (90.4%) | 41,564 (91.2%) | 8,790,075 (89.1%) | 93,798 (89.6%) |
| **Geography** |  |  |  |  |  |  |
| Rural | 2,479,688 (14.7%) | 29,056 (16.1%) | 616,941 (14.6%) | 7,069 (15.5%) | 1,340,756 (13.6%) | 15,499 (14.8%) |
| **Income Quintile** |  |  |  |  |  |  |
| 1st quintile | 2,982,780 (17.6%) | 33,814 (18.8%) | 736,716 (17.4%) | 8,612 (18.9%) | 1,842,945 (18.7%) | 20,940 (20.0%) |
| 2nd quintile | 3,335,222 (19.7%) | 36,536 (20.3%) | 835,276 (19.8%) | 8,847 (19.4%) | 2,007,802 (20.4%) | 21,849 (20.9%) |
| 3rd quintile | 3,351,614 (19.8%) | 35,237 (19.6%) | 838,012 (19.8%) | 9,043 (19.8%) | 1,997,302 (20.3%) | 21,266 (20.3%) |
| 4th quintile | 3,507,494 (20.7%) | 36,756 (20.4%) | 882,844 (20.9%) | 9,421 (20.7%) | 1,913,979 (19.4%) | 19,838 (19.0%) |
| 5th quintile | 3,702,063 (21.9%) | 37,314 (20.7%) | 921,278 (21.8%) | 9,521 (20.9%) | 2,084,008 (21.1%) | 20,613 (19.7%) |
| **Education Quintile** |  |  |  |  |  |  |
| 1st quintile | 3,075,278 (18.2%) | 34,957 (19.4%) | 762,687 (18.0%) | 8,779 (19.3%) | 1,706,122 (17.3%) | 19,391 (18.5%) |
| 2nd quintile | 3,385,511 (20.0%) | 37,128 (20.6%) | 841,692 (19.9%) | 9,481 (20.8%) | 1,937,907 (19.7%) | 21,469 (20.5%) |
| 3rd quintile | 3,493,804 (20.6%) | 36,945 (20.5%) | 870,402 (20.6%) | 9,190 (20.2%) | 2,030,870 (20.6%) | 21,454 (20.5%) |
| 4th quintile | 3,500,217 (20.7%) | 36,902 (20.5%) | 877,965 (20.8%) | 9,299 (20.4%) | 2,084,940 (21.1%) | 21,550 (20.6%) |
| 5th quintile | 3,357,516 (19.8%) | 33,022 (18.3%) | 844,839 (20.0%) | 8,485 (18.6%) | 2,034,100 (20.6%) | 20,045 (19.1%) |
| **Comorbidities** |  |  |  |  |  |  |
| AMI* | 709,309 (4.19%) | 9,667 (5.37%) | 177,912 (4.21%) | 2,507 (5.50%) | 416,924 (4.23%) | 5,696 (5.44%) |
| Arrhythmia | 1,530,096 (9.04%) | 18,798 (10.4%) | 382,872 (9.06%) | 4,891 (10.7%) | 914,621 (9.27%) | 10,990 (10.5%) |
| Arthritis | 354,214 (2.09%) | 4,455 (2.47%) | 89,539 (2.12%) | 1,113 (2.44%) | 225,636 (2.29%) | 2,719 (2.60%) |
| Asthma | 2,034,989 (12.0%) | 26,649 (14.8%) | 508,111 (12.0%) | 7,093 (15.6%) | 1,246,159 (12.6%) | 16,445 (15.7%) |
| Cancer | 2,198,985 (13.0%) | 25,044 (13.9%) | 546,554 (12.9%) | 6,210 (13.6%) | 1,317,439 (13.4%) | 14,662 (14.0%) |
| Chronic Heart Failure | 838,537 (4.95%) | 14,327 (7.95%) | 210,965 (4.99%) | 3,672 (8.06%) | 483,842 (4.91%) | 7,974 (7.62%) |
| Colitis | 133,502 (0.79%) | 1,575 (0.87%) | 32,232 (0.76%) | 389 (0.85%) | 88,715 (0.90%) | 1,018 (0.97%) |
| COPD** | 2,910,066 (17.2%) | 40,751 (22.6%) | 726,976 (17.2%) | 10,396 (22.8%) | 1,726,397 (17.5%) | 23,837 (22.8%) |
| Coronary Heart Disease | 3,510,361 (20.7%) | 43,579 (24.2%) | 879,995 (20.8%) | 11,068 (24.3%) | 1,899,361 (19.3%) | 23,359 (22.3%) |
| Dementia | 344,161 (2.03%) | 4,454 (2.47%) | 86,853 (2.06%) | 1,158 (2.54%) | 206,300 (2.09%) | 2,722 (2.60%) |
| Diabetes | 4,394,754 (26.0%) | 52,155 (29.0%) | 1,095,489 (25.9%) | 13,022 (28.6%) | 2,660,592 (27.0%) | 30,929 (29.5%) |
| Hypertension | 10,494,913 (62.0%) | 116,393 (64.6%) | 2,622,675 (62.1%) | 29,352 (64.4%) | 6,033,618 (61.2%) | 67,044 (64.0%) |
| Mental disorder*** | 3,410,850 (20.2%) | 40,032 (22.2%) | 851,249 (20.1%) | 10,117 (22.2%) | 2,244,412 (22.8%) | 26,407 (25.2%) |
| Mood disorder | 7,786,874 (46.0%) | 85,908 (47.7%) | 1,943,700 (46.0%) | 21,771 (47.8%) | 4,801,635 (48.7%) | 52,944 (50.6%) |
| Osteoarthritis | 10,244,732 (60.5%) | 111,020 (61.6%) | 2,553,893 (60.4%) | 28,336 (62.2%) | 6,131,185 (62.2%) | 66,621 (63.6%) |
| Osteoporosis | 1,791,824 (10.6%) | 18,620 (10.3%) | 450,339 (10.7%) | 4,656 (10.2%) | 1,032,043 (10.5%) | 10,525 (10.1%) |
| Renal Failure | 727,558 (4.30%) | 10,545 (5.85%) | 180,987 (4.28%) | 2,698 (5.92%) | 489,046 (4.96%) | 7,004 (6.69%) |
| Stroke | 965,743 (5.71%) | 12,601 (6.99%) | 239,406 (5.67%) | 3,122 (6.85%) | 547,368 (5.55%) | 7,026 (6.71%) |
| Total (median) | 3 ([2, 4]) | 3 ([2, 5]) | 3 ([2, 4]) | 3 ([2, 5]) | 3 ([2, 5]) | 3 ([2, 5]) |

\* Acute myocardial infarction
\*\* Chronic obstructive pulmonary disease
\*\*\* History of mental health-related visit, excluding dementia, deliberate self-harm codes, and mood disorder codes.

**Figure 1.**
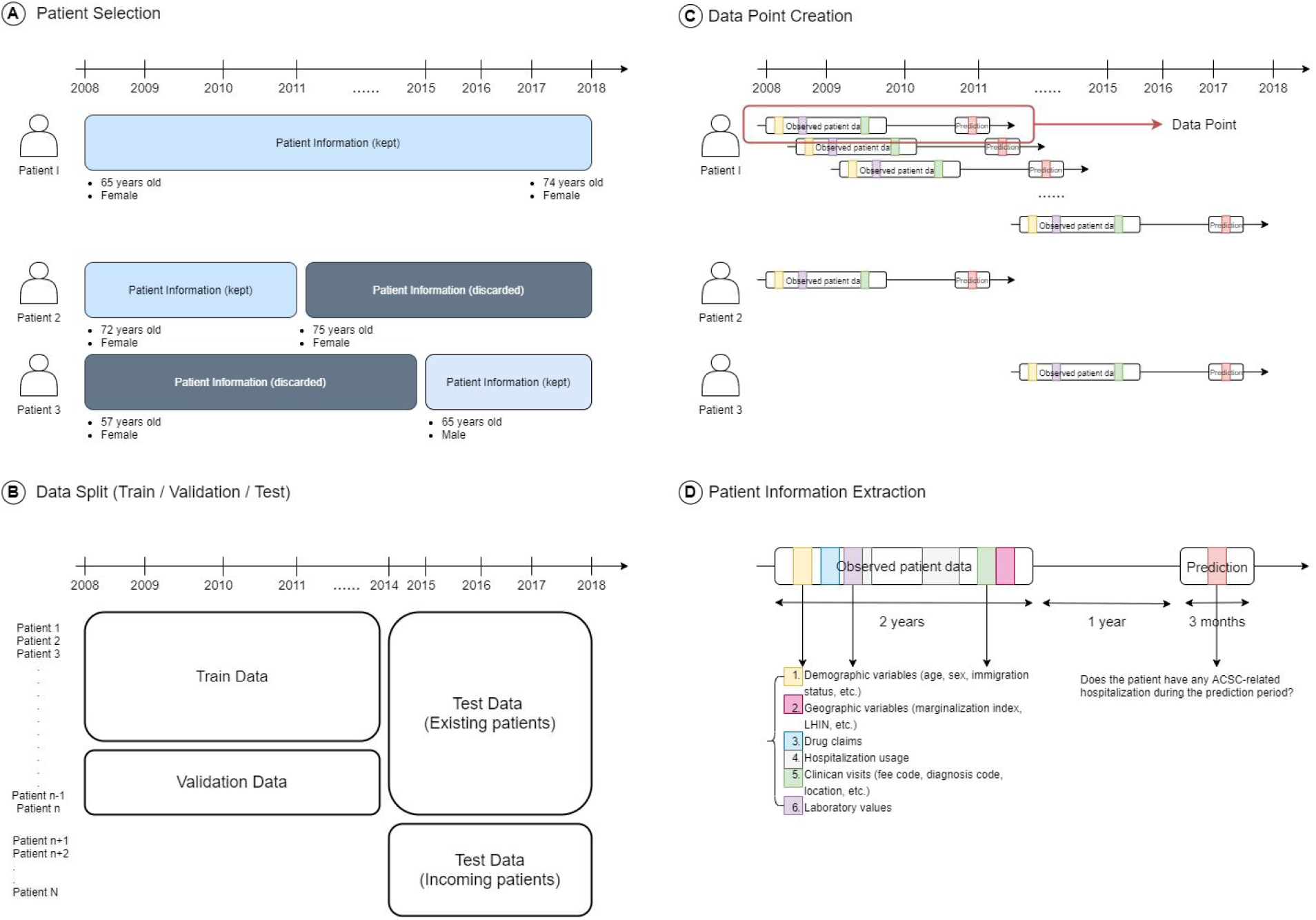
Overview of the Data Preparation. Panel A shows the patient selection process. For a given patient, only the information when they are between 65 and 74 years old was kept and the rest was discarded. This was done for the whole study period (between 2008 and 2017, included). After patients were selected, as shown in panel B, we split our data based on patients and study period. We trained and validated our prediction model with data between 2008 and 2014, on two different sets of patients. Then, we tested the model on two types of data: data of the patients who were already used for training and validation between 2014 and 2018, and data of the patients who entered the cohort in 2014 and onwards. The latter consisted of “young” patients between 65 and 66 years old at the beginning of the observation period. Panel C then shows the construction of instances for the eligible patients - each instance consisted of 2 years of observation window, 1 year of buffer where we didn’t have any information about the patient, and 3 months of target window. This *instance* was therefore a summary of the patient health information. Note that one patient could generate multiple instances. The first instance had an observation window from January 2008 to December 2009, a buffer from January 2010 to December 2010, and a target window from January 2011 to March 2011, all included. The last instance had an observation window from October 2014 to September 2016, a buffer from October 2016 to September 2017, and a target window from October 2017 to December 2017, all included. Instances for the same patient did not share any time period of the target window with each other, meaning each patient could have a maximum of 28 instances with non-overlapping target widows. This was often not the case due to the exclusion criteria and the individuals not fitting the age group of 65 to 74 years old anymore. Finally, panel D shows the types of information that was extracted from each patient.

### Study Design

As shown in Figure 1C, each patient’s timeline of interactions with the healthcare system was broken down into *instances*. Each instance comprised three components: two years of patient history (*observation window*), one year during which we did not extract any information (*buffer*), and a three-month period where we assess the risk of hospitalization due to an ACSC (*target window*). We used two years of patient history to collect enough information about the patient and calculate predictor variables. The three-month target window was selected due to the update frequency of administrative data at ICES, which also takes place every three months.

We split our cohort into three different sets (see Figure 1B). The training and validation sets had distinct patients, which ensures that we tested the ability of the model to generalize well on unseen patients. Both sets included target windows from January 2011 to December 2015. As for the test set, it included all instances of patients that have a valid target window between January 2016 and December 2017. Therefore, two types of patient groups are included in this test set: those from the training and validation sets who still qualify at a future period of time, and patients who were under 65 years old prior to 2016 but who qualify for the test time period. The model performance on the test set thus indicated how well the model generalizes to patients out-of-time and to new patients (i.e. in-domain, in-distribution generalization).

### Inclusion and Exclusion Criteria

Our cohort included all residents of Ontario between 65 and 74 years old during the study period. Anyone who became 65 during any given time of the study period was included in the study cohort, e.g., those who turned 65 in January 2017 were included in our test set, as shown in Figure 1. We excluded instances of patients who died or ended all interaction with the healthcare system before the end of the observation window. This was to ensure that we excluded patients who were by definition known to not have an outcome.

### Outcome of Interest

The Canadian Institute for Health Information (CIHI) defines age-standardized acute care hospitalization rate for conditions where appropriate ambulatory care is thought to prevent or reduce the need for admission to hospital [8]. There are seven different types of avoidable hospitalization-related ACSCs as per the CIHI definition: epilepsy, chronic obstructive pulmonary disease (COPD), asthma, diabetes, heart failure (HF), hypertension, and angina. For each hospitalization or ambulatory usage, we determined if the primary diagnosis code was in the list of CIHI ACSC diagnosis codes. We then aggregated all codes to determine whether or not an instance contained any hospitalization related to an ACSC from the above list during the target window, in which case we labelled the instance as a positive data point (i.e. our predictions are not cause-specific). The list of the ACSC diagnosis codes used as well as additional rules for defining an ACSC can be found in Supplementary material S3.

### Feature Preparation

The features that were extracted from the data sources included patients’ demographic and geographical information, drug prescription history, chronic conditions, clinician visits, hospital usage, laboratory results, as well as past history of ACSCs. In order to select the most important features for the model and reduce the likelihood of overfitting, we took a forward feature selection-based approach [28] to only select a small subset from thousands of available features that would ensure the performance of the model closely matches that of the model using all features. We ended with 140 features in total. Details of the feature preparation as well as the full list of the features are outlined in Supplementary material S4.

### Model Development and Evaluation

Our prediction model is a gradient boosting decision tree-based model, optimized using the XGBoost library in Python [29,30]. These models have been empirically shown as computationally efficient and high-performance algorithms for tabular data through a number of machine learning competitions as well as in machine learning for health papers [31]. These models are able to select and combine heterogeneous features from multiple data sources that are often uncorrelated, and are thus efficient when both categorical and continuous variables with different value ranges are present in the data. Moreover, they can implicitly handle missing values without imputation. Therefore, we did not impute missing data. We undersampled the negative instances of the training data by a factor of 8, to reduce class imbalance [32]. Validation and the test sets were left untouched to ensure an accurate evaluation of the model performance. The details of the model’s hyperparameters as well as its performance comparison against a baseline logistic regression model are presented in Supplementary material S5 and S6.

We evaluated the performance of the model on the held-out test set. We first checked the distribution of the predictions of our model after calibrating the probabilities to account for undersampling [33]. Model performance was then measured in terms of Area Under the Receiver Operating Curve (AUC), a widely used metric for risk prediction tasks [33,34]. Since the model was trained on highly imbalanced data, we also calculated the Area Under Precision-Recall Curve (AUPRC) to focus on the probability of correctly detecting patients with the highest risk of ACSC [35]. To assess the variation of model performance on different risk groups, we compared the model predictions on multiple subgroups created from the test set. These groups comprised different age, sex, immigration, and socioeconomic groups obtained using data across datasets (e.g. RPDB, CENSUS, ON-MARG in Supplementary table S1). We tested the model on patient groups split based on the number of interactions they had with the healthcare system as well.

Based on our model, we also computed Shapley Additive Explanations (SHAP) values, the weighted average of marginal contributions [36], to identify features that contribute the most to model predictions. We used a randomly selected pool of 50,000 instances to generate these values.

### Geospatial Variation

We finally examined the performance of our model in capturing geospatial variation of ACSC incidence rates in Ontario. Ontario has 14 Local Health Integrated Networks (LHINs), responsible for coordinating and funding local healthcare to improve resources access and patient experience [37]. The LHINs are further divided into 74 sub-Local Health Integrated Networks (sub-LHINs), and each of these aims at identifying health care needs and priorities within the region. We chose sub-LHINs to represent the subdivisions of the province. Following the predictions of our model, we computed and plotted the average risk of ACSCs for patients in each sub-LHIN.

### Patient and Public Involvement

No patients with ACSCs or members of the public at-large were involved in the conceptualization, analysis, or the write-up of this study. However, the public at-large is involved at ICES through the form of a Public Advisory Council which provides input into decisions made on how research is conducted using the individual-level personal health information collected. We plan to disseminate the knowledge gained through this study with the use of press releases and presentations on the value of population health planning, for both ACSCs and more broadly, tailored to general public audiences.

## Results

Starting from the initial cohort of 4,520,076 patients aged 50 years or older as of 2008, we excluded 1,873,139 patients who did not meet inclusion criteria due to age restrictions. From the remaining 2,646,937 patients, 752,505 were removed for their absence of interaction with the healthcare system, and 40,316 patients had a death date prior to the end of the first observation window. After the exclusion criteria, a total of 1,854,116 patients were selected, resulting in 31,010,869 instances. Among them, 1,237,507 patients with 16,921,175 instances were used for training, 309,380 patients with 4,227,660 instances for validation and 1,375,277 patients yielding 9,862,034 instances for testing. The ACSC incidence rates among instances were around 1.1% for all sets. Descriptive statistics for key demographic, socio-economic, and chronic illness variables are presented in Table 1.

### Model Performance

The model achieved an average AUC of 80.5 (range 80.4-80.5), when evaluated on all instances in the test set. Figure 2 shows the overall calibration curve of the model as well as model evaluation on important subgroups of the population. The calibration curve of Figure 2a. has 20 bins of equal data size, and shows the average risk of ACSC predicted by the model versus the actual ACSC-related hospitalization rate. The inset on the right side of the graph shows that the model is slightly overpredicting the risk.

**Figure 2.**
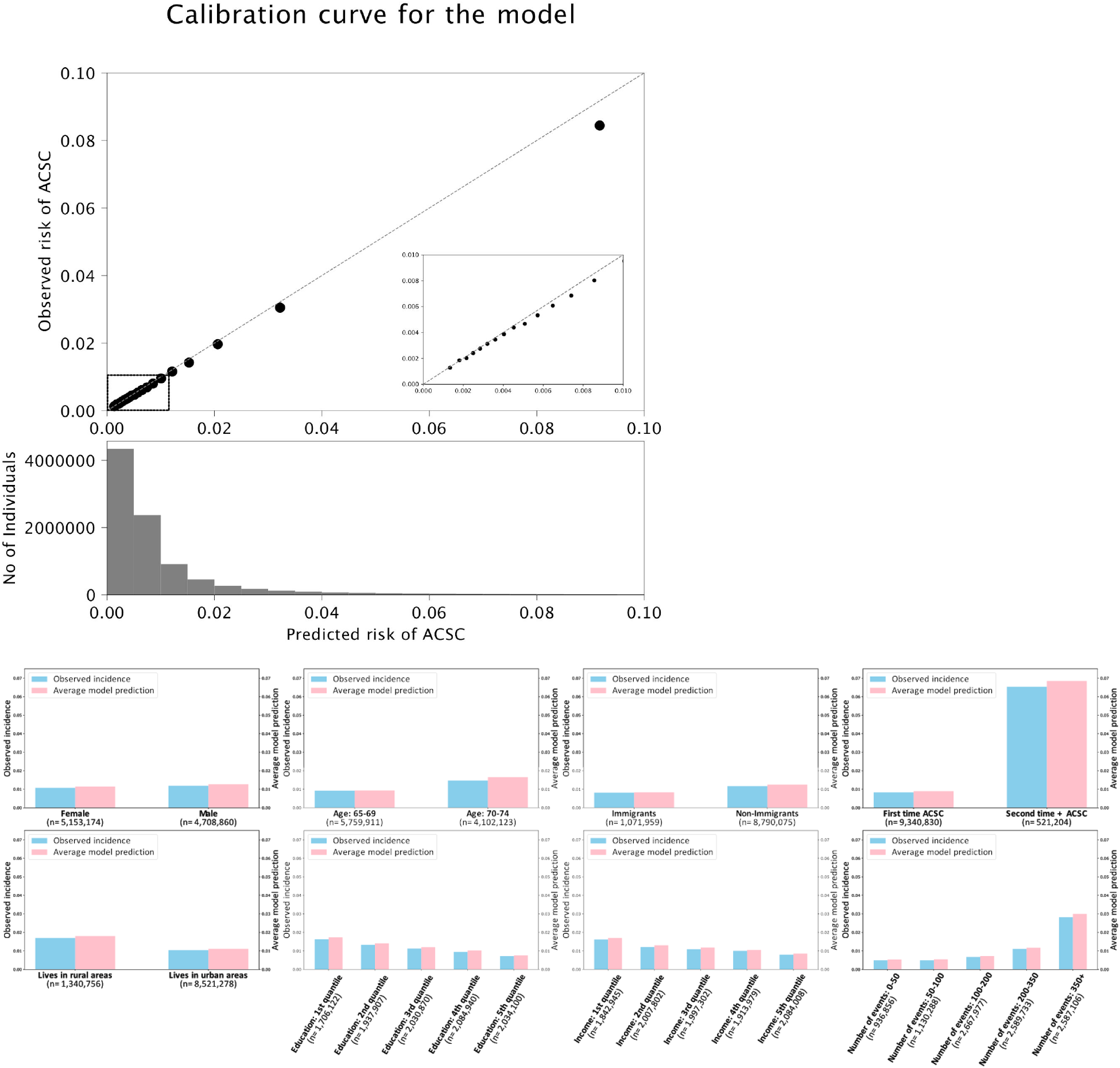
**a**. Calibration curve. **b**. Model evaluation on major subgroups of the population. The incidence rates are shown in blue and the average model prediction in pink. The subgroup sizes are displayed on the x-axis along with the subgroup types. For education and income quintiles, higher index refers to higher education level and income respectively, in the area a given patient lives in. The number of events refers to the number of any interaction a given patient had with the healthcare system - clinician visits, hospitalization, ambulatory usage, lab tests, and drug prescriptions.

Figure 2b. shows the average prediction of the model compared to the real incidence rate of ACSCs for different subgroups of the population. As in the calibration curve, the model slightly overpredicts the risk but accurately captures the variation in ACSC hospitalization risk across different subgroups (e.g. higher risk of hospitalization in a subcohort with a higher age or a lower income index). Ranking the predictions made by the model, we found that the top 5% of test set instances predicted by the model as “high-risk” covered 37.4% of instances who actually present an ACSC-related hospitalization during the target window period. The top 1% and 10% of test set instances covered 15.2% and 50.8% of total positive data points respectively. Main characteristics of these risk groups are described in Supplementary Figure S7.

### Feature Contribution

Figure 3 shows 20 features with the highest Shapley values. It is clear that the model is leveraging features across datasets, as the top features span demographics, geography, chronic conditions, prescriptions, and interactions with the healthcare system. The AUC of the model when trained on individual datasets as well as different combinations of the subsets (see Supplementary Figure S8) shows the impact of combining all features to achieve the best prediction performance. The previous number of ambulatory care visits, time since last ambulatory care visit, as well as presence of ACSC-related hospitalizations during the observation window were predictive of future ACSC-related hospitalizations. Several features related to medication prescription during the observation period were also found to be predictive of ACSC-related hospitalizations including the number of selective beta2-adrenergic agonists, absence of prescriptions for patients in long-term care facilities, and number of statin prescriptions. Geographical features such as latitude and rural residence were also predictive of our outcome. As shown in Supplementary Figure S9, the model predictions are influenced by different features depending on whether or not a given individual already has an ACSC-related hospitalization. Having a precedent was found to the contribution of future ACSC hospitalizations and the related features appear as the top supporting features.

**Figure 3.**
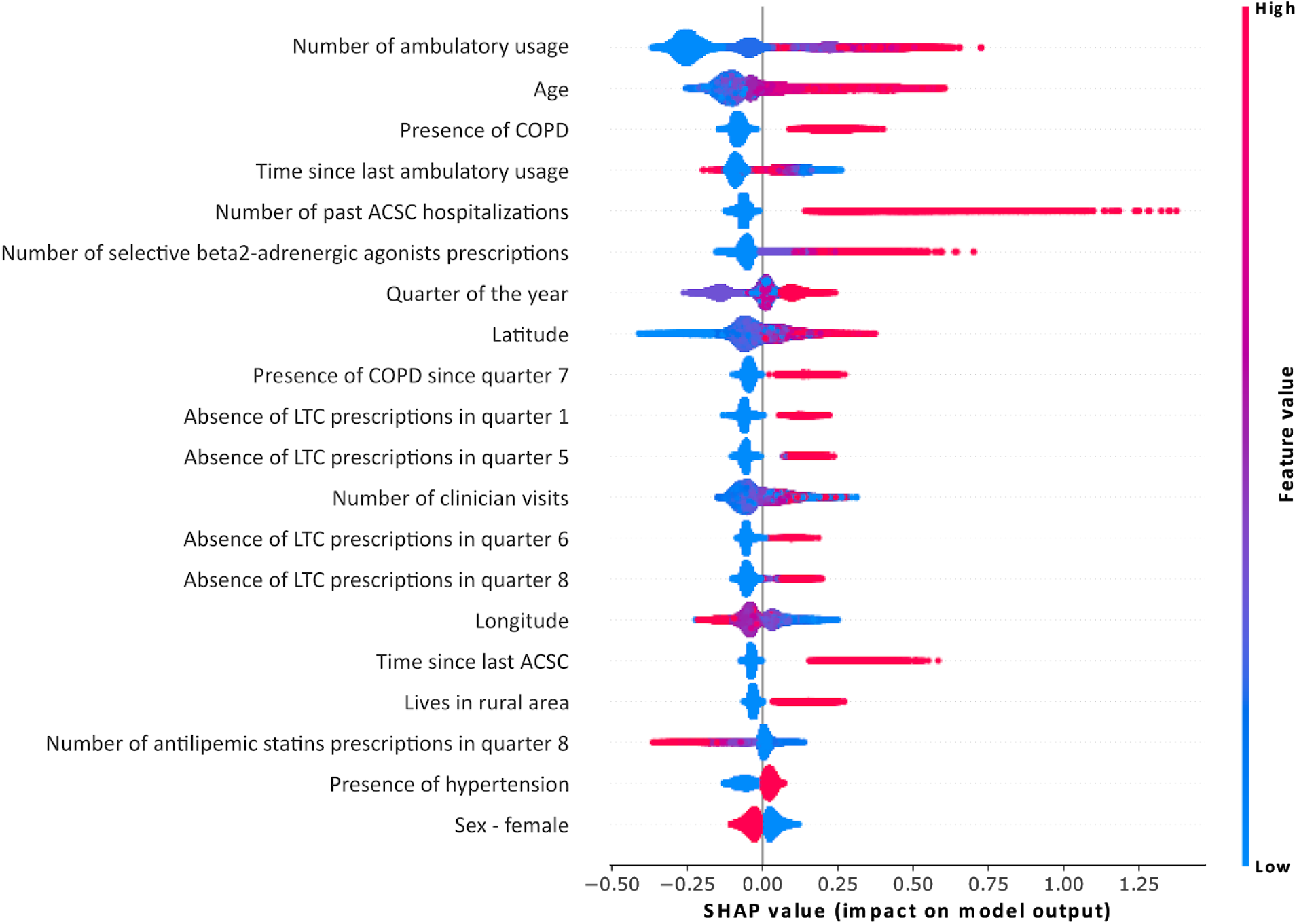
Global feature importance. Shapley values were generated using 50,000 random samples from the test set. Multiple runs using different samples showed the same ordering of feature contribution.

### Geospatial Variation

Figure 4 shows the geospatial variation of ACSC incidence rates in Ontario. Figure 4A (map in red) is the distribution of ACSC-related hospitalizations in different sub-LHINs of Ontario, weighted by the population size in each sub-LHIN. Figure 4B (map in blue) shows the distribution of ACSC risks predicted by the model. Our model is able to capture the geospatial heterogeneity of ACSC risk, in rural regions (e.g. Northwestern Ontario) as well as in urban and populous areas (e.g. Southern Ontario).

**Figure 4.**
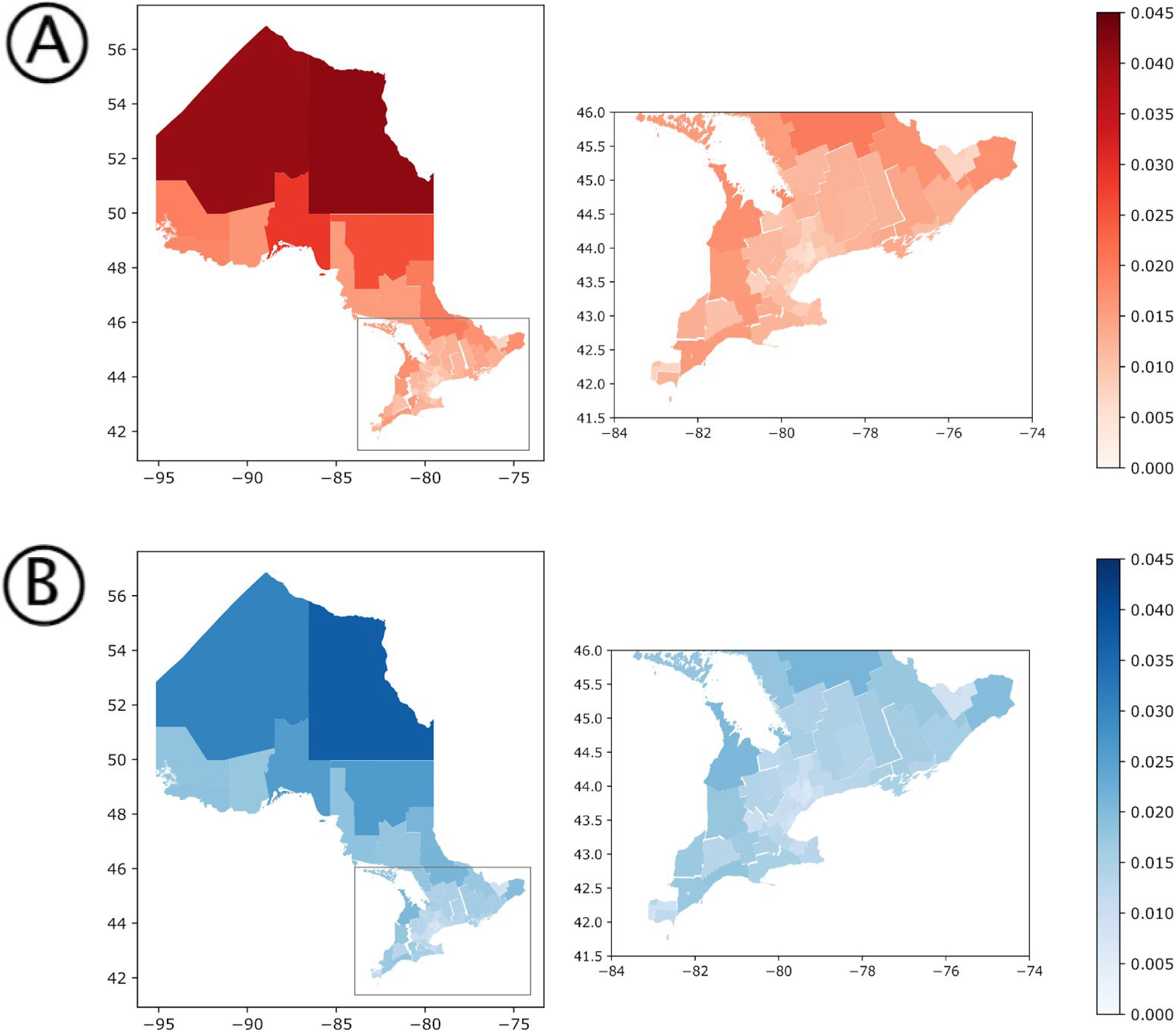
Distribution of ACSC risks predicted by the model compared to the actual variation in ACSC incidence rates in the province, in 2017. A) shows the incidence rates of ACSC by sub-LHIN, normalized by the population size of the corresponding sub-LHIN. For B), we computed the predictions of ACSC with our model for all patients and mapped them for different sub-LHINs. The model (in B) captures the normalized distribution of these patients (in A).

## Discussion

The goal of this work was to use AHD to develop a single, large-scale, machine learning model to predict hospitalizations due to ACSCs in a cohort of 1.85 million older adults in Ontario, Canada. This model is designed for population health planning and health resource allocation versus individual health decision-making. Our model accurately predicted the one-year risk for ACSCs with a high AUC and the prediction values are well-calibrated. The model leverages a variety of features across multiple administrative health databases to make its predictions, including outpatient visits, demographic information as well as past drug prescriptions. The presence of comorbidities such as COPD, hypertension, or chronic heart failure as well as geography were important features in the model which is consistent with previously identified risk factors in Canada. [10] Shapley values were provided for model transparency but should not be interpreted as causal risk factors. Our model is able to also capture geographic variations in risk across Ontario in both rural and urban settings.

### Strengths and Weaknesses in Relation to Other Studies

There have been numerous studies that predict the risk of emergency admissions in community-dwelling adults, some of which focus on those that are potentially avoidable [38]. A 2014 systematic review of 18 models that used either administrative or clinical data found that performance, as measured by the c-statistic, ranged between 0.63-0.83. Three of these studies focused on ACSCs [39,40,41]. The first is an Italian study that used AHD to identify adult patients at risk of hospitalization or death within a year who may be candidates for care through a ‘patient-centred Medical Home’ program [40]. We used similar sets of features, however, we were able to incorporate additional demographic and socioeconomic variables. Notably, we only created one single model for use in the entire population in order to facilitate the ease of application at the population level, while achieving similar model performance in terms of discrimination and calibration. The second study involved predicting the one-year risk of hospitalizations from ACSCs in a cohort from the Veteran’s Health Administration in the US [39]. An advantage of their work was the inclusion of individual-level socioeconomic variables. However, their cohort was much more homogenous than ours and designed for individual patient care of a very specific segment of the population. Such a model would not be applicable for population health planning or for use on routinely collected data where these individual determinants are not available. Finally, the Sussex Predictor of Key Events (SPOKE) model leveraged a neural network architecture with 1000 features across diverse health datasets to predict admission risk due to a range of chronic diseases for every individual in Sussex [41]. While it achieved a slightly higher AUC (0.82) than our model, its complex architecture and use of almost ten times the amount of features present serious challenges for model implementation and increases the likelihood of overfitting.

### Meaning of the Study to Clinicians and Policy Makers

With our model, we are able to assess the one-year risk of hospitalizations due to ACSCs by aggregating individual patient risks at a sub-regional level on a quarterly basis. It is important to see the ACSC rates not simply as a marker of access to primary care, but as an indicator for how resources could be allocated and optimized in a health system by better addressing the needs of specific regions and patient groups. In Ontario, it has been demonstrated that ambulatory care services for cardiovascular disease were actually provided more frequently in regions with lower rates of cardiovascular events (i.e. myocardial infarction, stroke, or cardiovascular-related death) [42]. Therefore, a model such as the one developed in this work is a valuable tool for health systems planners to inform health care delivery models and allocate community-based interventions over space and time with a focus on health equity [42,43]. Such reallocations are thought to be impactful, as it has been shown that readmission due to certain conditions, such as heart failure, can be reduced by the efficient and timely allocation of inpatient and outpatient counselling and monitoring efforts [44].

### Strengths and Weaknesses of the Study

Our study has several strengths. Our cohort is large and is constructed from a highly diverse population. We developed a model that uses a wide range of features to predict the risk of hospitalization for ACSCs, which is an indicator of the effectiveness of the healthcare system indicator. The model is designed in a way it can be deployed in a real-world setting at relatively low cost, using available administrative health data as well as area-level data that capture concepts related to the social determinants of health. It also allows risk assessment at different aggregation levels for geography and over time. Given that population-risk scores are known to be biased [45], we conducted a rigorous calibration evaluation not just globally, but in subgroups as well. However, our work also has some important limitations. A challenge lies in the focus of ACSC as an outcome, which has been the focus of some debate regarding the extent of its preventable nature [46]. Furthermore, the definition of ACSCs relies on diagnosis codes that are often considered to be a partial way of patient condition assessment [47]. It varies across different jurisdictions, even though the set of conditions used in this study is common in most definitions [5,7,8,46]. The lack of access to certain indicators (e.g. smoking, BMI, etc.) that are known to be important determinants of ACSC risks [10,39] is expected to bottleneck model performance and emphasizes the difficulty of assessing an individual’s risk without fully observing their health and behavioural status [48].

### Future Directions

While our study focused solely on AHD, it is theoretically possible to link this information with data captured in inpatient EMRs such that individual clinicians can use this information to direct specific care or treatments. The EMR would also facilitate the capture of individual clinical or risk behaviour features that are known to contribute ACSC-related hospitalization risk and thus may also increase model performance. This is a promising area of future work. Moreover, there is uncertainty around what interventions are most effective at reducing rates of hospitalizations in high-risk populations. For example, trials of care coordination programs have demonstrated mixed results for various chronic diseases [49,50]. Designing and validating these interventions across the population remains an important area for future work.

## Conclusion

In this work, we demonstrate that the development and validation of a single, large-scale machine learning model to predict the 1-year risk of hospitalization from a series of ambulatory-care sensitive conditions is feasible in a large and diverse cohort of seniors using administrative health data. Such a model has the potential to reduce the burden of hospitalizations from ambulatory care conditions by supporting the allocation of community-based interventions during population health planning and health resource allocation.

## Supporting information

TRIPOD Checklist

## Data Availability

The dataset for this study is held securely in coded form at ICES. While data sharing agreements prohibit ICES from making the dataset publicly available, access may be granted to those who meet pre- specified criteria for confidential access, available at www.ices.on.ca/DAS. The full dataset creation plan is available from the authors upon request.

## Summary Box

### What is already known on this topic

- Potentially preventable hospitalizations due to ambulatory care sensitive conditions (ACSCs) are a major focus for health system improvement
- There is a need for analytic tools that can run on routinely collected, population health data that can guide the deployment of interventions and the allocation of healthcare resources to reduce the burden of avoidable hospitalizations

### What this study adds

- This study outlines the development and validation of a novel machine learning model to predict future hospitalizations due to ACSCs that can be uniquely run on routinely collected administrative health data in a diverse population
- Our model has excellent discrimination, is well calibrated, and the top 5% of patients predicted as high risk represent 37% of ACSC-related hospitalizations
- Such a model may support the allocation of health system resources to reduce the burden of potentially avoidable hospitalizations

## Acknowledgements

We thank IMS Borgan Inc. for use of their Drug Information Database. This study was supported by ICES, which is funded by an annual grant from the Ontario Ministry of Health and Long-Term Care (MOHLTC). Parts of this material are based on data and information compiled and provided by MOHLTC, Canadian Institutes for Health Information (CIHI) and Immigration, Refugees, and Citizenship Canada (IRCC).

## Footnotes

### Contributors

SY and LR planned the study. TW prepared the cohort. SY analyzed the data with contributions from VH, JG, MR, MG, and MV. SY and VH wrote the first draft of the paper. All authors contributed to the critical revision of the manuscript for important intellectual content and approved the final version of the manuscript. The corresponding author attests that all listed authors meet authorship criteria and that no others meeting the criteria have been omitted. LR is the guarantor.

### Ethical Approval

ICES has obtained ethical approval (and repeats this review tri-annually) for its privacy and security policies, procedures, and practices. Each research project that is conducted at ICES is also subject to internal ethical review by the ICES Privacy and Compliance Office. ICES is a prescribed entity under section 45 of Ontario’s Personal Health Information Protection Act (PHIPA). Section 45 is the provision that enables analysis and compilation of statistical information related to the management, evaluation, and monitoring of, allocation of resources to, and planning for the health system. Section 45 authorizes health information custodians to disclose personal health information to a prescribed entity, like ICES, without consent for such purposes. Projects conducted wholly under section 45, by definition, do not require review by a Research Ethics Board. As a prescribed entity, ICES must submit to trio-annual review and approval of its privacy and security policies, procedures and practices by Ontario’s Information and Privacy Commissioner. These include policies, practices and procedures that require internal review and approval of every project by ICES’ Privacy and Compliance Office.

### Data Sharing

The dataset for this study is held securely in coded form at ICES. While data sharing agreements prohibit ICES from making the dataset publicly available, access may be granted to those who meet pre-specified criteria for confidential access, available at www.ices.on.ca/DAS. The full dataset creation plan is available from the authors upon request. The data for this study was prepared with custom code from ICES using the SAS Enterprise v6.1 software. This data was later analyzed with custom code from Layer 6 AI in the Java 8 and Python 3.6 programming languages. The analytic code is available from the authors upon request, understanding that the computer programs may rely upon coding templates or macros that are unique to ICES and this data and thus may require modification.

### Transparency Declaration

The lead and corresponding authors affirm that the manuscript is an honest, accurate, and transparent account of the study being reported; that no important aspects of the study have been omitted; and that any discrepancies from the study as planned (and, if relevant, registered) have been explained.

### Dissemination to participants and related patient and public communities

We plan to disseminate the knowledge gained through this study with the use of press releases with lay abstracts and presentations on the value of population health planning, for both ACSCs and more broadly, tailored to general public audiences.

### Provenance and peer review

Not commissioned; externally peer reviewed.

