## Supplementary material for "Predicting hospitalizations related to ambulatory care sensitive conditions with machine learning for population health planning: derivation and validation cohort study"

### S1 & S2. Datasets used

| Dataset | Description |
|---|---|
| **RPDB (Registered Persons Database)** | The RPDB provides basic demographic information (age, sex, location of residence, date of birth, and date of death for deceased individuals) for those issued an Ontario health insurance number. The RPDB also indicates the time periods for which an individual was eligible to receive publicly funded health insurance benef ts and the best known postal code for each registrant on July 1st of each year. |
| **IRCC (Immigration, Refugees and Citizenship Canada's Permanent Resident Database)** | The Ontario portion of the IRCC Permanent Resident Database includes immigration application records for people who initially applied to land in Ontario since 1985. The dataset contains permanent residents' demographic information such as country of citizenship, level of education, mother tongue, and landing date. New immigrants who are currently residing in Ontario but originally landed in another province are not captured in this dataset. |
| **ON-MARG (Ontario Marginalization Index)** | ONMARG is a geographically (census) based index developed to quantify the degree of marginalization occurring across the province of Ontario. It is comprised of four major dimensions thought to underlie the construct of marginalization: residential instability, material deprivation, dependency, and ethnic concentration. The dataset contains census divisions (CD), census tracts (CT), census subdivisions (CSD), consolidated municipal service manager areas (CMSM), public health units (PHU), local health integration networks (LHIN), sub-LHINs, and dissemination areas (DA). |
| **CENSUS** | The 2006, 2011, and 2016 Canadian census were used to capture information on area-level income and education. |
| **Multimorbidity Dataset** | This dataset was created combining multiple datasets such as OHIP, DAD, and ICES-derived chronic disease cohorts. It summarizes the diagnosis date of 18 chronic conditions for each patient. For more detail on the def nition used to identify each chronic condition, see Table S2. |
| **DAD (Discharge Abstract Database)** | The DAD is compiled by the Canadian Institute for Health Information and contains administrative, clinical (diagnoses and procedures/interventions), demographic, and administrative information for all admissions to acute care hospitals, rehab, chronic, and day surgery institutions in Ontario. At ICES, consecutive DAD records are linked together to form 'episodes of care' among the hospitals to which patients have been transferred after their initial admission. |
| **NACRS (National Ambulatory Care Reporting System)** | The NACRS is compiled by the Canadian Institute for Health Information and contains administrative, clinical (diagnoses and procedures), demographic, and administrative information for all patient visits made to hospital- and community-based ambulatory care centres (emergency departments, day surgery units, hemodialysis units, and cancer care clinics). At ICES, NACRS records are linked with other data sources (DAD, OMHRS) to identify transitions to other care settings, such as inpatient acute care or psychiatric care. |
| **DIN (Druglist File)** | The DIN f le contains a near exhaustive list of drug identif cation numbers used in Canada from 1990 forward. Contains information on drug and product names (generic and trade names), subclass information, PCG codes, Drug strength, Route of Administration, f rst and last dispensing dates from ODB. |
| **ODB (Ontario Drug Benef t)** | The ODB database contains prescription medication claims for those covered under the provincial drug program, mainly: those aged 65 years and older, nursing home residents, patients receiving services under the Ontario Home Care program, those receiving social assistance, and residents eligible for specialized drug programs. Main data elements include drug identif er, quantity, days supplied, date dispensed, cost, and patient, pharmacy and physician identif ers. |
| **OHIP (Ontario Health Insurance Plan)** | The OHIP claims database contains information on inpatient and outpatient services provided to Ontario residents eligible for the province's publicly funded health insurance system by fee-for-service health care practitioners (primarily physicians) and "shadow billings" for those paid through non-fee-for-service payment plans. The main data elements include patient and physician identif ers (encrypted), code for service provided, date of service, associated diagnosis, and fee paid. |
| **OLIS (Ontario Laboratory Information System)** | OLIS is a province-wide integrated repository of patients' lab test orders and results. Lab tests and results related to Hemoglobin A1C carried out from 2008 to 2015 were included for the study. |

**Table S1**. Dataset Description. Details of the datasets used are presented. Descriptions were adapted from the ICES data dictionary: https://datadictionary.ices.on.ca/Applications/DataDictionary/Default.aspx

The list S2 of 18 chronic conditions that were used for the model input is attached in a separate document.

#### S3. Ambulatory Care Sensitive Conditions (ACSCs)

| Type of Criteria | Criteria | Details |
|---|---|---|
| **Inclusions** | Hospitalization for an ambulatory care sensitive condition is identif ed as any most responsible diagnosis code of: | - **Chronic obstructive pulmonary disease (COPD):** Any most responsible diagnosis (MRDx) code of J41, J42, J43, J44, J47, or MRDx of acute lower respiratory infection (J10.0, J11.0, J12–J16, J18, J20, J21, J22), only when a *secondary diagnosis** of J44 is also present.<br>**Secondary diagnosis* refers to a diagnosis other than the most responsible one.<br>- **Grand mal status and other epileptic convulsions:** G40, G41<br>- **Asthma:** J45.<br>- **Diabetes:** E10.0, E10.1, E10.63, E10.64, E10.9, E11.0, E11.1, E11.63, E11.64, E11.9, E13.0, E13.1, E13.63, E13.64, E13.9, E14.0, E14.1, E14.63, E14.64, E14.90.<br>- **Heart failure and pulmonary edema:** I50, J81.<br>- **Hypertension:** I10.0, I10.1, I11.<br>- **Angina:** I20, I23.82, I24.0, I24.8, I24.9. |
| | Admission to an acute care institution (Facility Type Code = 1). | |
| | Age at admission younger than 75. | |
| **Exclusions** | For heart failure and pulmonary edema, hypertension, and angina, exclude cases with cardiac procedures: | The full list of cardiac procedure codes for exclusion can be found in the CIHI Def nition [2].<br>Codes may be coded in any position. Procedures coded as abandoned after onset (Intervention Status Attribute = A) are excluded. |
| | Records with missing sex. | |
| | Records with discharge as death (Discharge Disposition Code = 07, 72*, 73*, 74*). | |
| | Newborn, stillbirth or cadaveric donor records (Admission Category Code = N, R or S). | |

**Table S3**. Definition of Ambulatory Care Sensitive Conditions adapted from the criteria given by the Canadian Institute for Health Information [1]. All codes are in ICD-10-CA [2].

#### S4. Final selected features

The features that were extracted from the data sources include patients' demographic and geographical information, drug prescription history, chronic conditions, clinician visits, hospital usage, as well as past history of ACSC and laboratory results.

Demographic information included the age of the patient, their sex, immigration status, and the date of arrival in Canada and the country of birth if applicable. Geographical information not only consisted of the address of the patient at the three postal code digit level (also called as a *forward sortation area* of *FSA*), but also of the aggregation of different socioeconomic status measures at the FSA-level. This included quintiles of area-level education or income, as well as marginalization indices measuring material deprivation, residential instability, dependency, and ethnic concentration.

Chronic conditions of the patient examine the presence of 18 comorbidities such as hypertension, diabetes, chronic obstructive pulmonary disease, and mood disorder. Drug prescription history has the information of the quantity and the name of medications that were prescribed to a given patient. It also has the information of whether or not the medication was prescribed in a long-term care facility. It is important to note that this contains the information of the medications that were dispensed but we cannot know if they were actually taken by the patient. These records are nevertheless a good proxy for the condition of the patient as well as an indicator of whether or not the patient is exposed to polypharmacy, the concurrent use of more than five medications. The physician visits and hospital usage information contained the type of physician or hospital visit and the fee code related to the visit. Finally, from hospitalization data, information on the presence and type of ACSC-related hospitalization was extracted for each patient.

To control for the diversity of the patients, we set a threshold to the frequency of each feature to avoid processing very rare features values that are not generalizable- for instance, the drug class information was processed only if at least 25% of the patients were prescribed medications of the same class. All categorical values were one-hot encoded. Demographic and geographical features were prepared as fixed attributes of the patient at the time of observation window. The other features were aggregated at a quarterly-level to account for the characteristics of the datasets being updated every three months. We also aggregated the latter at the observation window-level to obtain global health status of the patient, such as the total number of prescriptions of drug class A or the time since the last ACSC-related hospitalization.

The total number of features reached 2,082 after the initial preparation. In order to select the most important features for the model as well as to ensure its generalizability, we took a greedy approach to select a small subset of features that would ensure the performance of the model to match that of the model using all features. Starting from a subset of 50 most contributing and geographic features we wanted to keep in, other features were added to the subset only if it led to a visible increase in model performance when evaluated on the validation set. At the end of the process, we ended with 140 features in total.

The complete list S4 of final features that were selected for the model is attached in a separate document.

#### S5. Model Development Specification

Given the low incidence rate of ACSCs (1.83% in the training set), we had a very imbalanced dataset and while training, we undersampled negative data points (no ACSC hospitalization in prediction window) by selecting only one out of 8 negative samples, and kept all positive data points (ACSC hospitalization in prediction window). The validation and testing sets were left untouched (i.e. were not undersampled). The final predictions made by the model were calibrated to account for undersampling [3]. The model was trained with the following hyperparameters: a learning rate of 0.05, a maximum tree depth of 10, and both the fraction of columns to be randomly sampled and the subsample ratio of columns for each split set at 0.7. The alpha, gamma and lambda values were 0.3, 0.1 and 0.5 [4]. These were selected after a hyperparameter grid search, consisting in fixing ranges and increments for given hyperparameters and testing all combinations of values to find the optimal one [5] .

#### S6. Model performance in comparison with Logistic Regression

We trained a Logistic Regression (LR), a model that is widely used in developing healthcare risk prediction models. The LR model was trained using the same features as the XGBoost model. As seen in Table S5, XGBoost model is able to predict the risk of ACSC-related hospitalizations with a higher AUC. While the XGBoost model is able to handle multiple types of variables without any feature engineering, LR requires feature normalization and all features were scaled between 0 and 1.

We compared the AUC value for the whole cohort of patients, where we saw a gain of 1.2 by using the XGBoost model. We reported the range of AUC as well as its average obtained by training the model 5 times with random restarts. We also compared the AUC values when looking at the “young” patients who newly qualified by turning 65 during the test set study period (target window between January 2016 and December 2017). This ensures that the algorithm will be able to correctly assess the risk of the patients who just turn 65 and are added to the patient group in the test set. XGBoost again shows an AUC gain of 1.4 compared to LR. We also compared the precision-recall curve of our XGBoost model to a logistic regression model. The rapid drop in precision is predictable due to the rarity of the ACSC-related outcomes.

| | All Patients | New Patients |
|---|---|---|
| **Logistic Regression** | 79.3 (79.2-79.5) | 78.4 (78.2-78.6) |
| **XGBoost** | 80.5 (80.4-80.5) | 79.8 (79.6-79.9) |

**Table S5.** AUC values for XGBoost and LR on all patients and newly added patients to the test set.

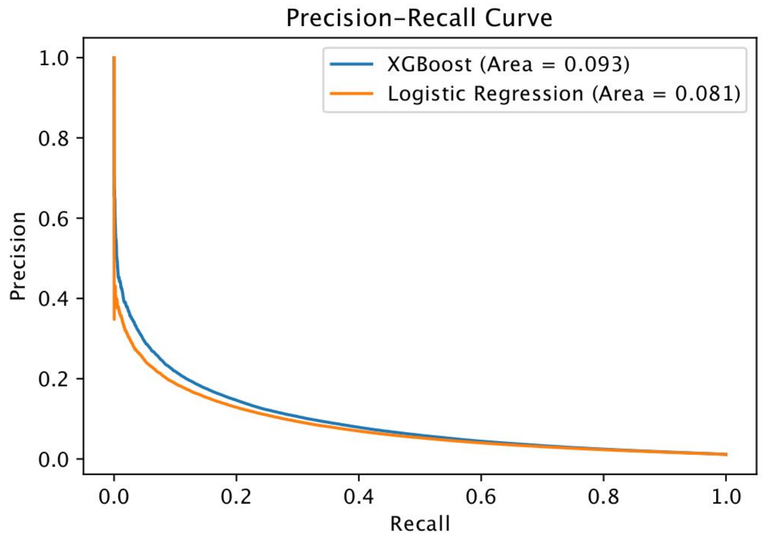


**Figure S6** Precision-Recall Curve comparing XGBoost model against Logistic Regression.

#### S7. Characteristics of different risk groups identified by the model

| | Top 1% | Top 5% | Top 10% | All patients |
|---|---|---|---|---|
| **Sex - Female (%)** | 50.2 | 48.7 | 48.7 | 52.3 |
| **Sex - Male (%)** | 49.8 | 51.3 | 51.3 | 47.7 |
| **Age (mean)** | 70.5 | 70.1 | 69.9 | 69.0 |
| **Immigrant (%)** | 3.61 | 4.46 | 4.92 | 10.9 |
| **Non-Immigrant (%)** | 96.4 | 95.5 | 95.1 | 89.1 |
| **No history of ACSC (%)** | 13.4 | 45.9 | 62.8 | 94.7 |
| **History of ACSC (%)** | 86.6 | 54.1 | 37.2 | 5.28 |
| **Lives in Rural Areas (%)** | 24.3 | 23.5 | 22.9 | 13.6 |
| **Lives in Urban Areas (%)** | 75.7 | 76.5 | 77.1 | 86.4 |
| **Education quantile (mean)** | 2.41 | 2.51 | 2.57 | 3.09 |
| **Income quantile (mean)** | 2.45 | 2.54 | 2.60 | 3.05 |
| **Number of events (median)** | 688 | 524 | 439 | 210 |

**Table S7.** Baseline characteristics comparison for patients in dif erent risk level groups, predicted by the model. For education and income quintiles, higher index refers to higher education level and income respectively, in the area a given patient lives in. The number of events refers to the number of any interaction a given patient had with the healthcare system - clinician visits, hospitalization, ambulatory usage, lab tests, and drug prescriptions.

#### S8. Model performance for different dataset types.

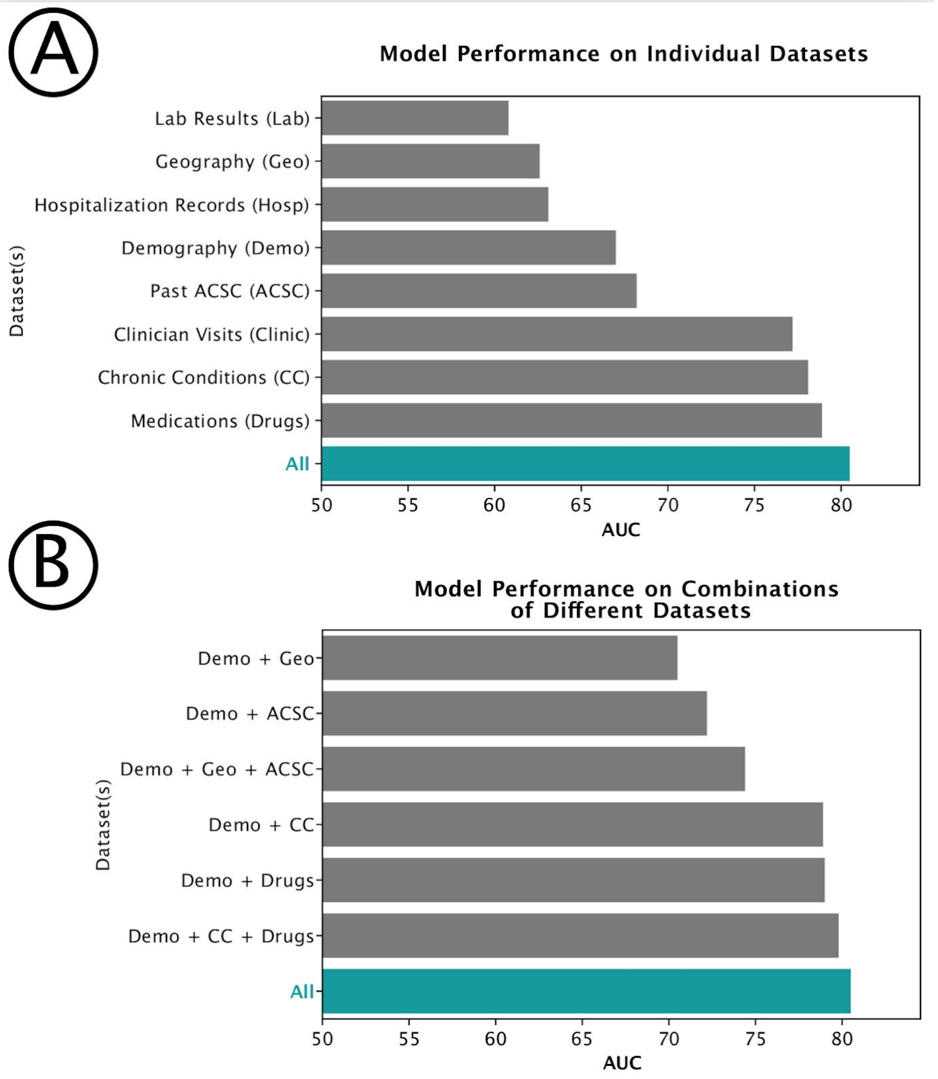


**Figure S8** : Model performance on different features of subsets.

#### S9. Feature contribution for different population subgroups

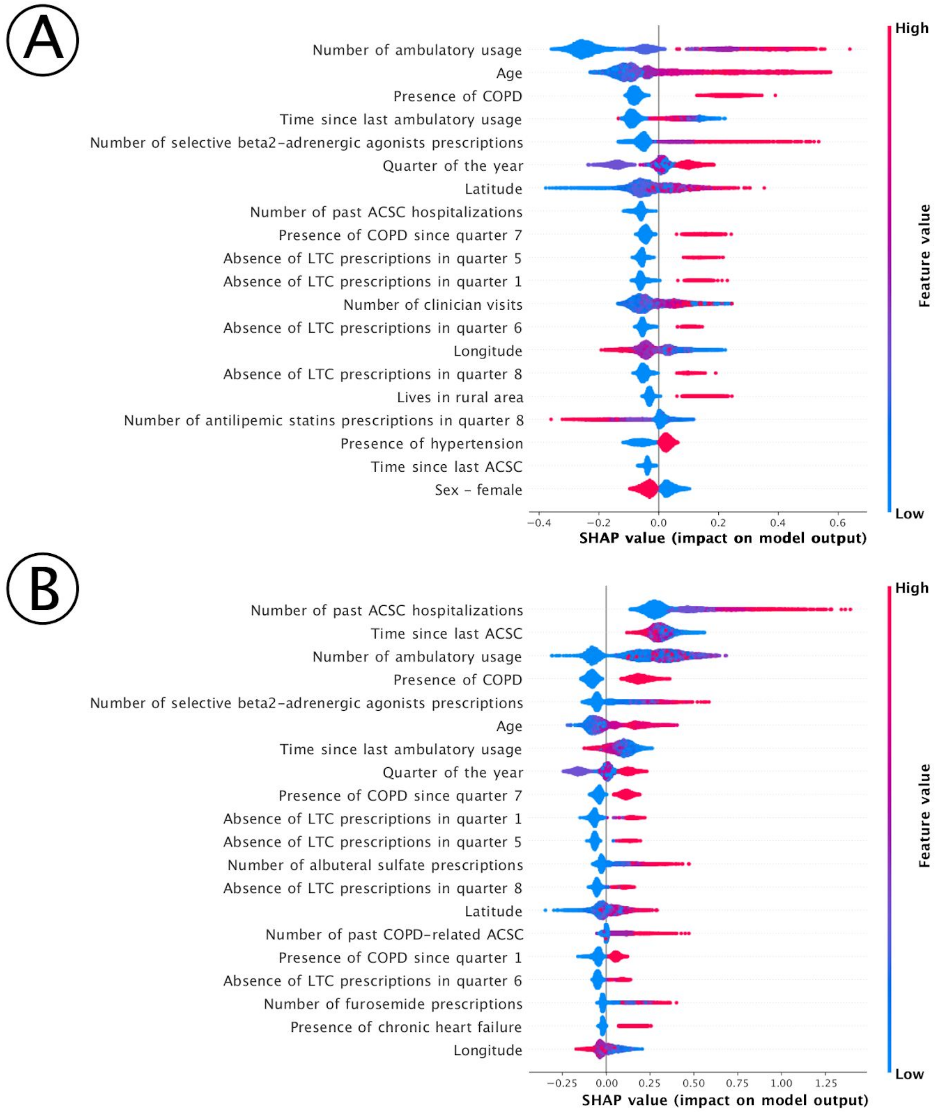


**Figure S9** A) Feature importance for patients who do not have a history of an ACSC-related hospitalization. B) Feature importance for patients who have a history of one or more ACSC-related hospitalizations.
