## Supplementary table S2 for "Predicting hospitalizations related to ambulatory care sensitive conditions with machine learning for population health planning: derivation and validation cohort study"

| **Chronic condition** | **Definition / Source** | **Lookback Window*** |
|---|---|---|
| Asthma | ICES-derived cohort: Ontario Asthma Database (ASTHMA) | 1992 |
| Cancer | Ontario Cancer Registry (OCR) | 1992 |
| Congestive Heart Failure | ICES-derived cohort: Ontario Congestive Heart Failure Database (CHF) | 1992 |
| Chronic Obstructive Pulmonary Disorder | ICES-derived cohort: Ontario Chronic Obstructive Pulmonary Disease (COPD) | 1992 |
| Diabetes | ICES-derived cohort: Ontario Diabetes Database (ODD) | 1992 |
| Acute Myocardial Infarction | One hospitalization in DAD using the ICD codes (ICD9: 410, ICD10: I21) | 1988 (DAD) |
| Rheumatoid Arthritis | ICES-derived cohort: Ontario Rheumatoid Arthritis Database (ORAD) | 1992 |
| Osteo- and other Arthritis | (1) One hospitalization in DAD; OR (2) Two or more OHIP physician billing claim within a two-year period using the ICD codes:<br>ICD9/OHIP: 715, 710, 711, 716, 718, 720, 727, 728, 729, 739, 274<br>ICD10: M00-M03, M07, M10, M11-M14, M20-M25, M30-M36, M65-M79, M15-M19 | 1991 (OHIP)<br>1988 (DAD) |
| Crohn's Or Colitis | ICES-derived cohort: Ontario Chron's and Colitis Cohort Database (OCCC) | 1992 |
| Cardiac Arrhythmia | (1) One hospitalization in DAD; OR (2) Two or more OHIP physician billing claim within a two-year period using the ICD codes:<br>ICD9 /OHIP: 427.3 (DAD) / 427 (OHIP)<br>ICD 10: I48.0, I48.1 | 1991 (OHIP)<br>1988 (DAD) |
| Hypertension | ICES-derived cohort: Ontario Hypertension Database (HYPER) | 1992 |
| Chronic Coronary Syndrome | (1) One hospitalization in DAD; OR (2) Two or more OHIP physician billing claim within a two-year period using the ICD codes:<br>ICD 9/OHIP: 411-414<br>ICD-10: I20, I22-I25 | 1991 (OHIP)<br>1988 (DAD) |
| Stroke (Excluding TIA) | (1) One hospitalization in DAD; OR (2) Two or more OHIP physician billing claim within a two-year period using the ICD codes:<br>Any hospital admission with the following dx codes:<br>ICD-9: 430, 431, 432, 434, 436<br>ICD-10: 160 (excl 160.8), 161, I62, 163 (excl 163.6), 164 | 1991 (OHIP)<br>1988 (DAD) |
| Osteoporosis | ICES-derived cohort: (1) One hospitalization in DAD; OR (2) Two or more OHIP physician billing claim within a two-year period using the ICD codes:<br>ICD9/OHIP: 733<br>ICD10: M81, M82 | 1991 (OHIP)<br>1988 (DAD) |

| Chronic condition | Definition / Source | Lookback Window* |
|---|---|---|
| Mood Disorder (History of Mental Health-related Visit) | (1) One hospitalization in DAD/OMHRS; OR (2) Two or more OHIP physician billing claim within a two-year period using the ICD codes:<br>From DAD var DX10CODE1 with any of the following ICD-10-CA codes:<br>From OMHRS:<br>- If var AXIS1_DSM4CODE_DISCH1 complete (i.e,. listed diagnosis from below present) use AXIS1_DSM4CODE_DISCH1<br>- No, use PROVDX1<br>- Exclude OMHRS admissions if AXIS1_DSM4CODE_DISCH1 in: (290.x OR 294.x). If AXIS1_DSM4CODE_DISCH1 missing, exclude if PROVDX1=2<br>- Include visits/admissions with suspect diagnoses (suspect = T).<br>ICD9/OHIP: 311, 309, 300, 296<br>ICD10: F30—F34 (excl. F340), F38—F42, F431, F432, F438, F44, F450, F451, F452, F48, F530, F680, F930, F99 | 1991 (OHIP)<br>1988 (DAD)<br>2005 (OMHRS) |
| Other Mental Health Disorder (History of Mental Health-related Visit)<br>- Note: This excludes dementia, deliberate self-harm codes, and mood disorder codes. | (1) One hospitalization in DAD/OMHRS; OR (2) Two or more OHIP physician billing claim within a two-year period using the ICD codes:<br>From DAD var DX10CODE1 with any of the following ICD-10-CA codes:<br>From OMHRS:<br>- If var AXIS1_DSM4CODE_DISCH1 complete (i.e,. listed diagnosis from below present) use AXIS1_DSM4CODE_DISCH1<br>- No, use PROVDX1<br>- Exclude OMHRS admissions if AXIS1_DSM4CODE_DISCH1 in: (290.x OR 294.x). If AXIS1_DSM4CODE_DISCH1 missing, exclude if PROVDX1=2<br>- Include visits/admissions with suspect diagnoses (suspect = T).<br>ICD9/OHIP: 291, 292, 295, 297, 298, 299, 301, 302, 303, 304, 305, 306, 307, 313, 314, 315, 319<br>ICD10: F04, F050, F058, F059, F060, F061, F062, F063, F064, F07, F08, F10, F11, F12, F13, F14, F15, F16, F17, F18, F19, F20 , F21, F22, F23, F24, F25, F26 F27 F28, F29, F340, F35, F36, F37, F430, F439, F453, F454, F458, F46, F47, F49, F50, F51, F52, F531, F538, F539, F54, F55, F56, F57, F58, F59, F60, F61, F62, F63, F64, F65, F66, F67, F681, F688, F69, F70, F71, F72, F73, F74 F75 F76 F77 F78, F79, F80, F81, F82, F83, F84, F85 F86 F87 F88, F89, F90, F91, F92, F931, F932, F933, F938, F939, F94, F95, F96, F97, F98 | 1991 (OHIP)<br>1988 (DAD)<br>2005 (OMHRS) |

| **Chronic condition** | **Definition / Source** | **Lookback Window*** |
|---|---|---|
| Dementia | (1) One hospitalization in DAD; OR (2) Two or more OHIP physician billing claim within a two-year period using the ICD codes:<br>ICD9/OHIP: 290, 331 (OHIP) / (DAD: 046.1, 290, 294, 331.0, 331.1, 331.5, 331.82)<br>ICD10: F00, F01, F02, F03, G30 | 1991 (OHIP)<br>1988 (DAD) |
| Renal Failure | (1) One hospitalization in DAD; OR (2) Two or more OHIP physician billing claim within a two-year period using the ICD codes:<br>ICD9/OHIP: 403,404,584,585,586,v451<br>ICD 10: N17, N18, N19, T82.4, Z49.2, Z99.2 | 1991 (OHIP)<br>1988 (DAD) |

**Table S2**. List of 18 chronic conditions used for the Multimorbidity Dataset.
$*$ Lookback window refers to the time window used to extract the history of chronic conditions.
