## Supplementary table S4 for "Predicting hospitalizations related to ambulatory care sensitive conditions with machine learning for population health planning: derivation and validation cohort study"

| Feature Name | Dataset | Description |
|---|---|---|
| Age | RPDB | Age of the patient at the end of the observation window |
| Sex (female) | RPDB | Sex of the patient (binary) |
| Quarter of the year | - | Quarter of the year at the end of the observation window. |
| LHIN (1-14) | ON-MARG | Local Health Integrated Network the patient lives in. Takes on a binary value for each LHIN from 1 to 14. (14 features in total) |
| Living in rural areas | ON-MARG | Binary indicator of whether or not a patient lives in a rural area. |
| Marginalization Index - Dependency (1st quintile - 5th quintile) | ON-MARG | Index to quantify the dependency level of the neighborhood the patient lives in. Takes on a binary value for each index level from 1 to 5. Higher index indicates lower degree of marginalization. (5 features in total) |
| Marginalization Index - Ethnicity (1st quintile - 5th quintile) | ON-MARG | Index to quantify the ethnic marginalization of the neighborhood the patient lives in. Takes on a binary value for each index level from 1 to 5. Higher index indicates lower degree of marginalization. (5 features in total) |
| Marginalization Index - Instability (1st quintile - 5th quintile) | ON-MARG | Index to quantify the instability level of the neighborhood the patient lives in. Takes on a binary value for each index level from 1 to 5. Higher index indicates lower degree of marginalization. (5 features in total) |
| Marginalization Index - Deprivation (1st quintile - 5th quintile) | ON-MARG | Index to quantify the deprivation level of the neighborhood the patient lives in. Takes on a binary value for each index level from 1 to 5. Higher index indicates lower degree of marginalization. (5 features in total) |
| Income (1st quintile - 5th quintile) | ON-MARG | Index to quantify the income level of the neighborhood the patient lives in. Takes on a binary value for each index level from 1 to 5. Higher index indicates higher income. (5 features in total) |
| Education (1st quintile - 5th quintile) | ON-MARG | Index to quantify the education level of the neighborhood the patient lives in. Takes on a binary value for each index level from 1 to 5. Higher index indicates higher education level. (5 features in total) |
| Latitude | ON-MARG | Value in decimal degrees to a precision of 2 decimal places ($\sim 1km$). |
| Longitude | ON-MARG | Value in decimal degrees to a precision of 2 decimal places ($\sim 1km$). |
| Presence of arrythmia | MMB Macro | Presence of arrythmia throughout the observation window. |
| Presence of asthma | MMB Macro | Presence of asthma throughout the observation window. |
| Presence of asthma since quarter 7 | MMB Macro | Presence of asthma starting from the 7th quarter of the observation window (at the latest). |

| Feature Name | Dataset | Description |
|---|---|---|
| Presence of chronic heart failure | MMB Macro | Presence of chronic heart failure throughout the observation window. |
| Presence of chronic heart failure since quarter 1 | MMB Macro | Presence of chronic heart failure starting from the 1st quarter of the observation window (at the latest). |
| Presence of chronic heart failure since quarter 7 | MMB Macro | Presence of chronic heart failure starting from the 7th quarter of the observation window (at the latest). |
| Presence of COPD | MMB Macro | Presence of chronic obstructive pulmonary disease throughout the observation window. |
| Presence of COPD since quarter 1 | MMB Macro | Presence of COPD starting from the 1st quarter of the observation window (at the latest). |
| Presence of COPD since quarter 3 | MMB Macro | Presence of COPD starting from the 3rd quarter of the observation window (at the latest). |
| Presence of COPD since quarter 4 | MMB Macro | Presence of COPD starting from the 4th quarter of the observation window (at the latest). |
| Presence of COPD since quarter 5 | MMB Macro | Presence of COPD starting from the 5th quarter of the observation window (at the latest). |
| Presence of COPD since quarter 6 | MMB Macro | Presence of COPD starting from the 6th quarter of the observation window (at the latest). |
| Presence of COPD since quarter 7 | MMB Macro | Presence of COPD starting from the 7th quarter of the observation window (at the latest). |
| Presence of coronary disease | MMB Macro | Presence of coronary disease throughout the observation window. |
| Presence of coronary disease since quarter 1 | MMB Macro | Presence of coronary disease starting from the 1st quarter of the observation window (at the latest). |
| Presence of coronary disease since quarter 2 | MMB Macro | Presence of coronary disease starting from the 2nd quarter of the observation window (at the latest). |
| Presence of coronary disease since quarter 7 | MMB Macro | Presence of coronary disease starting from the 7th quarter of the observation window (at the latest). |
| Presence of diabetes | MMB Macro | Presence of diabetes throughout the observation window. |
| Presence of diabetes since quarter 1 | MMB Macro | Presence of diabetes starting from the 1st quarter of the observation window (at the latest). |
| Presence of diabetes since quarter 2 | MMB Macro | Presence of diabetes starting from the 2nd quarter of the observation window (at the latest). |
| Presence of diabetes since quarter 3 | MMB Macro | Presence of diabetes starting from the 3rd quarter of the observation window (at the latest). |
| Presence of diabetes since quarter 4 | MMB Macro | Presence of diabetes starting from the 4th quarter of the observation window (at the latest). |
| Presence of diabetes since quarter 5 | MMB Macro | Presence of diabetes starting from the 5th quarter of the observation window (at the latest). |
| Presence of diabetes since quarter 7 | MMB Macro | Presence of diabetes starting from the 7th quarter of the observation window (at the latest). |
| Presence of hypertension | MMB Macro | Presence of hypertension throughout the observation window. |
| Presence of hypertension since quarter 7 | MMB Macro | Presence of hypertension starting from the 7th quarter of the observation window (at the latest). |

| Feature Name | Dataset | Description |
|---|---|---|
| Presence of mental disease | MMB Macro | Presence of mental disease throughout the observation window. |
| Presence of mental disease since quarter 1 | MMB Macro | Presence of mental disease starting from the 1st quarter of the observation window (at the latest). |
| Presence of mental disease since quarter 7 | MMB Macro | Presence of mental disease starting from the 7th quarter of the observation window (at the latest). |
| Presence of mood disorder | MMB Macro | Presence of mood disorder throughout the observation window. |
| Presence of renal failure | MMB Macro | Presence of renal failure throughout the observation window. |
| Presence of stroke since quarter 1 | MMB Macro | Presence of stroke starting from the 1st quarter of the observation window (at the latest). |
| Number of ambulatory usage | NACRS | Number of ambulatory usage of the patient over the observation window |
| Time since last ambulatory usage | NACRS | If there is, time since the last ambulatory usage of the patient over the observation window |
| Presence of ACSC hospitalization | DAD, NACRS | Binary indicator of the presence of any ACSC-related hospitalization over the observation window |
| Time since last ACSC | DAD, NACRS | If there is, time since the last ACSC-related hospitalization over the observation window |
| Number of clinician visits | OHIP | Number of clinician visits of the patient over the observation window |
| Number of selective beta2-adrenergic agonists prescriptions | ODB | Number of selective beta2-adrenergic agonists prescriptions over the observation window |
| Time since last selective beta2-adrenergic agonists prescriptions | ODB | If there is, time since last selective beta2-adrenergic agonists prescription over the observation window |
| Number of beta-blockers prescriptions | ODB | Number of beta-blockers prescriptions over the observation window |
| Time since last beta-blockers prescriptions | ODB | If there is, time since last beta-blockers prescription over the observation window |
| Number of furosemide prescriptions | ODB | Number of furosemide prescriptions over the observation window |
| Number of albuterol sulfate prescriptions | ODB | Number of albuterol sulfate prescriptions over the observation window |
| Number of antilipemic statins prescriptions in quarter 8 | ODB | Number of antilipemic statins prescriptions in the 8th quarter of the observation window |
| Time since last lab test | OLIS | If there is, time since last lab test the patient had over the observation window |
| Absence of LTC prescriptions in quarter 1 | ODB | Binary indicator of LTC prescriptions over the first quarter of the observation window |
| Clinician visit feecode in quarter 8 - chest radiology | OHIP | Number of clinician visits related to chest radiology in the 8th quarter of the observation window |
| Number of calcium blockers prescriptions | ODB | Number of calcium blockers prescriptions over the observation window |

| Feature Name | Dataset | Description |
|---|---|---|
| Number of lab tests | OLIS | Total number lab tests the patient had over the observation window |
| Number of hospitalizations | DAD | Total number of hospitalizations the patient had over the observation window |
| Clinician visit location in quarter 4 - office | OHIP | Number of visits to a clinician in office during the 4th quarter of the observation window |
| Number of antilipemic statins prescriptions in quarter 5 | ODB | Number of antilipemic statins prescriptions in the 5th quarter of the observation window |
| Absence of LTC prescriptions in quarter 5 | ODB | Binary indicator of LTC prescriptions over the 5th quarter of the observation window |
| Absence of LTC prescriptions in quarter 6 | ODB | Binary indicator of LTC prescriptions over the 6th quarter of the observation window |
| Absence of LTC prescriptions in quarter 8 | ODB | Binary indicator of LTC prescriptions over the 8th quarter of the observation window |
| Number of antilipemic statins prescriptions in quarter 7 | ODB | Number of antilipemic statins prescriptions in the 7th quarter of the observation window |
| Time since last furosemide prescription | ODB | If there is, the last time the patient was prescribed with furosemide during the observation window |
| Clinician visit feecode in quarter 7 - chest radiology | OHIP | Number of clinician visits related to chest radiology in the 7th quarter of the observation window |
| Clinician visit feecode in quarter 6 - chest radiology | OHIP | Number of clinician visits related to chest radiology in the 6th quarter of the observation window |
| Clinician visit feecode in quarter 5 - chest radiology | OHIP | Number of clinician visits related to chest radiology in the 5th quarter of the observation window |
| Time since last albuterol sulfate prescription | ODB | If there is, the last time the patient was prescribed with albuterol sulfate during the observation window |
| Number of macrolides prescriptions | ODB | Number of macrolides prescriptions over the observation window |
| Time since last calcium blocker prescription | ODB | If there is, the last time the patient was prescribed with calcium blocker during the observation window |
| Clinician visit feecode in quarter 4 - chest radiology | OHIP | Number of clinician visits related to chest radiology in the 4th quarter of the observation window |
| Clinician visit feecode in quarter 2 - chest radiology | OHIP | Number of clinician visits related to chest radiology in the 2nd quarter of the observation window |
| Clinician visit location in quarter 2 - office | OHIP | Number of visits to a clinician in office during the 2nd quarter of the observation window |
| Clinician visit feecode in quarter 1 - chest radiology | OHIP | Number of clinician visits related to chest radiology in the 1st quarter of the observation window |
| Clinician specialty in quarter 8 - emergency medicine | OHIP | Number of visits to a clinician specialized in emergency medicine during the 8th quarter of the observation window |
| Number of non-steroidal anti-inflammatory prescriptions | ODB | Number of non-steroidal anti-inflammatory prescriptions during the observation window |
| Time since last antilipemic statins prescription | ODB | If there is, the last time the patient was prescribed with antilipemic statins during the observation window |

| Feature Name | Dataset | Description |
|---|---|---|
| Emergency visit in quarter 8 - no blood transfusion | NACRS | No blood transfusion during emergency visits in the 8th quarter during the observation window |
| Time since last clinician visit | OHIP | If there is, the last time the patient goes for a clinician visit over the observation window |
| Number of antilipemic statins prescriptions in quarter 6 | ODB | Number of antilipemic statins prescriptions in the 6th quarter of the observation window |
| Clinician visit location in quarter 6 - office | OHIP | Number of visits to a clinician in office during the 6th quarter of the observation window |
| Clinician specialty in quarter 7 - emergency medicine | OHIP | Number of visits to a clinician specialized in emergency medicine during the 7th quarter of the observation window |
| Clinician visit location in quarter 5 - office | OHIP | Number of visits to a clinician in office during the 5th quarter of the observation window |
| Presence of COPD-related ACSC | DAD, NACRS | Binary indicator of the presence of any COPD ACSC-related hospitalization over the observation window |
| Number of benzodiazepine prescriptions | ODB | Number of benzodiazepine prescriptions over the observation window |
| Number of fluoroquinolones prescriptions | ODB | Number of fluoroquinolones prescriptions over the observation window |
| Number of antilipemic statins prescriptions in quarter 3 | ODB | Number of antilipemic statins prescriptions in the 3rd quarter of the observation window |
| Number of antilipemic statins prescriptions in quarter 1 | ODB | Number of antilipemic statins prescriptions in the 1st quarter of the observation window |
| Clinician specialty in quarter 3 - internal medicine | OHIP | Number of visits to a clinician specialized in internal medicine during the 3rd quarter of the observation window |
| Time since last fluoroquinolones prescriptions | ODB | If there is, time since last fluoroquinolones prescription over the observation window |
| Clinician visit feecode in quarter 3 - chest radiology | OHIP | Number of clinician visits related to chest radiology in the 3rd quarter of the observation window |
| Time since last narcotics (opiate agonists) prescriptions | ODB | If there is, time since last narcotics (opiate agonists) prescription over the observation window |
| Time since last ACE inhibitors prescriptions | ODB | If there is, time since last ACE inhibitors prescription over the observation window |
| Number of ACE inhibitors prescriptions | ODB | Number of ACE inhibitors prescriptions over the observation window |
| Number of narcotics (opiate agonists) prescriptions | ODB | Number of narcotics (opiate agonists) prescriptions over the observation window |
| Clinician specialty in quarter 7 - dermatology | OHIP | Number of visits to a clinician specialized in dermatology during the 7th quarter of the observation window |
| Clinician specialty in quarter 6 - emergency medicine | OHIP | Number of visits to a clinician specialized in emergency medicine during the 6th quarter of the observation window |

**Table S4**. List of all features used in the XGBoost model and Linear Regression.
